# Evaluation of *S*/*F*_94_ as a proxy for COVID-19 severity

**DOI:** 10.1101/2022.09.25.22280081

**Authors:** Maaike C Swets, Steven Kerr, James Scott-Brown, Adam B Brown, Rishi Gupta, Jonathan E Millar, Enti Spata, Fiona McCurrach, Andrew D Bretherick, Annemarie Docherty, David Harrison, Kathy Rowan, Neil Young, ISARIC4C Investigators, Geert H Groeneveld, Jake Dunning, Jonathan S Nguyen-Van-Tam, Peter Openshaw, Peter W. Horby, Ewen Harrison, Natalie Staplin, Malcolm G Semple, Nazir Lone, J Kenneth Baillie

## Abstract

Optimising statistical power in early-stage trials and observational studies accelerates discovery and improves the reliability of results. Ideally, intermediate outcomes should be continuously distributed and lie on the causal pathway between an intervention and a definitive outcome such as mortality. In order to optimise power for an intermediate outcome in the RECOVERY trial, we devised and evaluated a modification to a simple, pragmatic measure of oxygenation function - the *S*_*a*_*O*_2_/*F*_*I*_*O*_2_ (S/F) ratio.

We demonstrate that, because of the ceiling effect in oxyhaemoglobin saturation, *S/F* ceases to reflect pulmonary oxygenation function at high values of *S*_*a*_*O*_2_. Using synthetic and real data, we found that the correlation of *S/F* with a gold standard (*P*_*a*_*O*_2_/*F*_*I*_*O*_2_, *P/F* ratio) improved substantially when measurements with *S*_*a*_*O*_2_ ≥ 0.94 are excluded (Spearman *r*, synthetic data: *S/F* : 0.31; *S/F*_94_: 0.85). We refer to this measure as *S/F*_94_.

In order to test the underlying assumptions and validity of *S/F*_94_ as a predictor of a definitive outcome (mortality), we collected an observational dataset including over 39,000 hospitalised patients with COVID-19 in the ISARIC4C study. We first demonstrated that *S/F*_94_ is predictive of mortality in COVID-19. We then compared the sample sizes required for trials using different outcome measures (*S/F*_94_, the WHO ordinal scale, sustained improvement at day 28 and mortality at day 28) ensuring comparable effect sizes. The smallest sample size was needed when *S/F*_94_ on day 5 was used as an outcome measure.

To facilitate future study design, we provide an online user interface to quantify real-world power for a range of outcomes and inclusion criteria, using a synthetic dataset retaining the population-level clinical associations in real data accrued in ISARIC4C https://isaric4c.net/endpoints.

We demonstrated that *S/F*_94_ is superior to *S/F* as a measure of pulmonary oxygenation function and is an effective intermediate outcome measure in COVID-19. It is a simple and non-invasive measurement, representative of disease severity and provides greater statistical power to detect treatment differences than other intermediate endpoints.

## Introduction

Therapeutic research in COVID-19 depends on efficient, accurate assessment of therapeutic candidates in early-stage clinical studies. Efficacy measures should be “clinically meaningful”^1^ endpoints such as the WHO ordinal scale.^2^ Intermediate endpoints for early phase trials, or severity measures for observational studies, must be closely related to the causal pathway from intervention to a key outcome such as mortality, must be modifiable by therapy, and ideally should have a continuous numerical distribution to improve statistical power.^3^

In COVID-19, intermediate endpoints such as the WHO ordinal scale, duration of hospitalisation, and viral load have been used widely.^4, 5^ Both the WHO ordinal scale and various alternative ordinal scales,^6, 7^ rely on a complex clinical measure - the level of respiratory support received by a patient - as an indicator of illness severity. Viral load is a valid outcome for antiviral therapy, but it has not been shown to correlate with mortality benefit, and is not directly relevant to the effect of anti-inflammatory treatments.^8–10^ In the RECOVERY trial, we identified a need for more powerful intermediate endpoints for early phase clinical trials.[citation to follow: MEDRXIV/2022/280285]

Impairment of lung oxygenation function indicates disease progression in COVID-19,^11^ and is strongly predictive of mortality.^12^ Importantly, in COVID-19, failure of lung oxygenation is likely to be mechanistically linked to death: patients at extreme risk of mortality^12^ have high survival rates if oxygenation is provided by extracorporeal membrane oxygenation (ECMO).^13^ Pulmonary oxygenation function, together with clinical decision-making and resource availability, determines movement between most of the stages of the WHO Ordinal Scale (WHO scale points 4-9).^2^ Oxygenation function is a key determinant of efficacy for immunosuppression with corticosteroids in COVID-19.^9^ It is likely that lung oxygenation function lies on the causal pathway between the SARS-CoV-2 infection and death for many hospitalised patients.

Peripheral oxygen saturation can be measured easily and non-invasively using a pulse oximeter (formally, arterial oxygen saturation measured by pulse oximetry, rather than direct measurement in blood, is *S*_*p*_*O*_2_). The ratio of *S*_*a*_*O*_2_ or *S*_*p*_*O*_2_ to inspired fraction of oxygen (*F*_*I*_*O*_2_), known as the *S/F* ratio, provides a continuous index of lung oxygenation function which can be calculated without an arterial blood sample. *S/F* correlates well with the most widely-used arterial blood-derived measure of oxygenation - *P/F* ratio (*P*_*a*_*O*_2_/*F*_*I*_*O*_2_).^14^ *S/F* under steady state conditions in humans can range from around 0.5 (severe oxygenation defect) to 4.8 (perfect oxygenation function). A major limitation of *S/F* is the ceiling effect: at high *S*_*a*_*O*_2_ values, *S*_*a*_*O*_2_ ceases to be dependent on lung oxygenation function, because the blood is close to maximally oxygenated.^15, 16^ A healthy patient with perfect lungs breathing 21% oxygen with *S*_*a*_*O*_2_= 0.99 would have *S/F* = 4.7, but the same patient breathing 100% oxygen would have *S/F* = 0.99.

In order to improve the accuracy of measurement of lung oxygenation, we limited the ceiling effect in prospective data by protocolising measurement of *S*_*a*_*O*_2_ to control high values, or in retrospective analyses by excluding values recorded with *S*_*a*_*O*_2_ above a given value. We first evaluated an optimal threshold using both synthetic and real data from arterial blood gas samples, concluding that *S*_*a*_*O*_2_ *<* 0.94 provides an optimal balance between safety and predictive validity. We defined the *S/F*_94_ measurement as: *S/F* measured when *S*_*a*_*O*_2_ is 0.94 or less, achieved by reducing *F*_*I*_*O*_2_ to a minimum of = 0.21 (the fraction of oxygen in ambient air). Since many patients receive oxygen through devices for which *F*_*I*_*O*_2_ is not accurately quantified (e.g. Hudson mask, nasal cannulae), prospective studies measuring *S/F*_94_ will require a protocolised modification of oxygen delivery devices which, in itself, is expected to improve accuracy of measurement (Appendix: Protocol). Here, we assess the predictive validity of this outcome measure and compare it to a range of alternative outcome measures.

## Results

### Relationship with gold standard oxygenation measure (*P/F*)

There is a consistent pattern in both synthetic (Figure 1) and real (Supplementary Figure 1) data: if no maximum cut-off value for *S*_*a*_*O*_2_ is used, spuriously low *S/F* values are seen in patients with good lung function, reflected in high *P/F* values (Figure 1a, Supplementary Figure 1a). This is due to the ceiling effect - *S*_*a*_*O*_2_ cannot rise above 100%. These misleading values are removed by excluding values with *S*_*a*_*O*_2_ above 94% (Figure 1b, Supplementary Figure 1b), which improves the correlation with the gold standard for both synthetic (Spearman *rS/F* : 0.40; *S/F*_94_: 0.85; Figure 1a) and real data (Spearman *rS/F* : 0.82; *S/F*_94_: 0.97, Supplementary Figure 1a).

**Figure 1:**
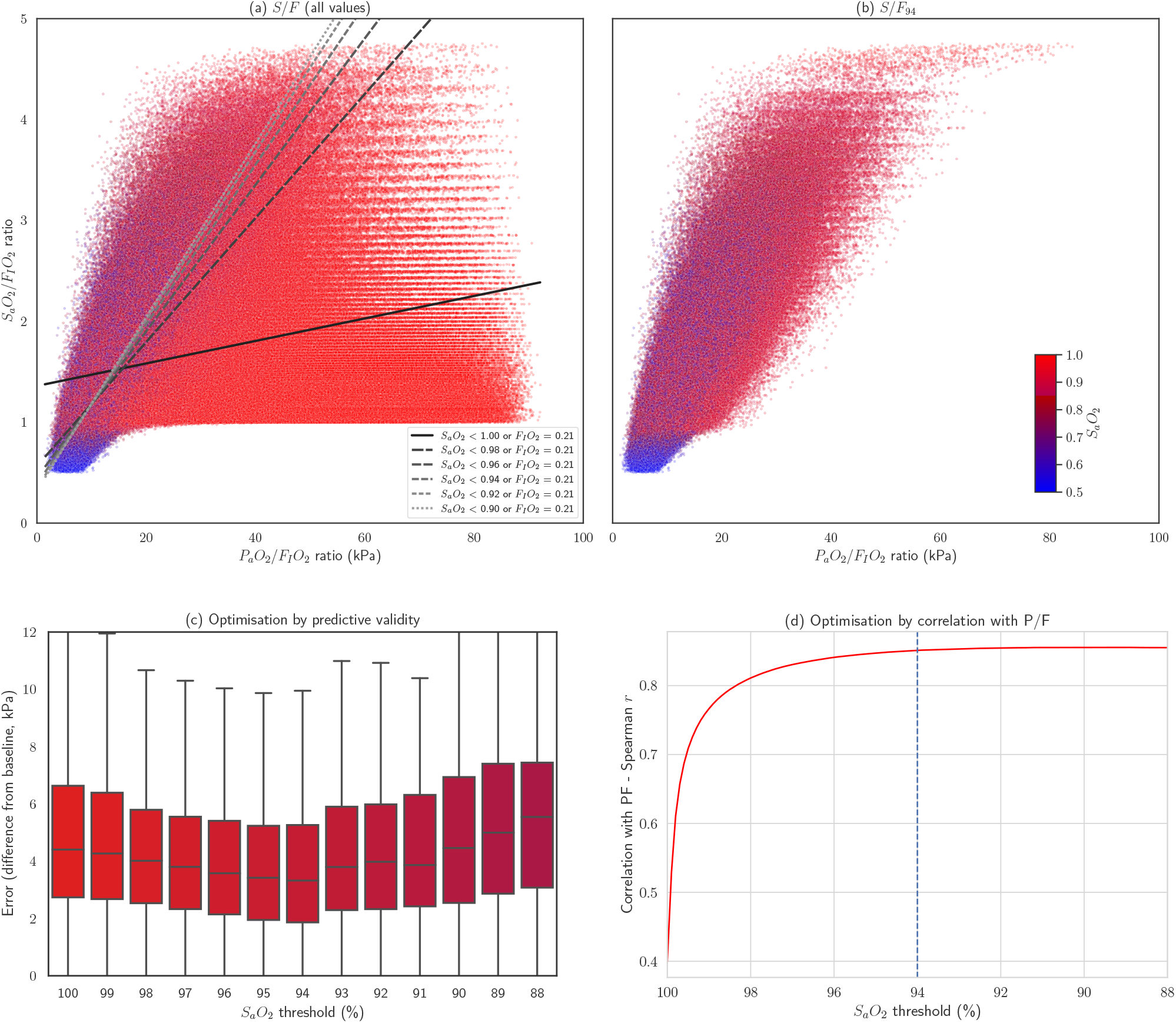
Comparison of *P/F* and *S/F* or *S/F*_94_ in synthetic data. (a,b) Scatterplots of *P/F* vs *S/F* individual measurements across a range of hypothetical physiological characteristics. Points are coloured according the *S*_*a*_*O*_2_ as shown in the colour scale. (a) including all values, showing linear regression of *S/F* against *P/F* in using different cut-off values for *S*_*a*_*O*_2_. Patients breathing air (*F*_*I*_*O*_2_=21%) were included in all bins. (b) including only values with *S*_*a*_*O*_2_ *<* 94% or *F*_*I*_*O*_2_ = 21% (c,d) Optimisation of cut-off value for *S*_*a*_*O*_2_ using predictive validity: the error in the prediction of a future *P*_*a*_*O*_2_, based on a previous one. (c) (d) change in correlation coefficient (Pearson’s *ρ*) as the threshold for inclusion is lowered from *S*_*a*_*O*_2_*<* 100% to *S*_*a*_*O*_2_*<* 80%.

### Predictive validity

In parallel, we assessed the predictive validity of *S/F* and *S/F*_94_. As in our previous work,^17^ we assert that if *S/F*_94_ is measuring true oxygenation function well, then it should be able to more accurately predict a future event: the *P*_*a*_*O*_2_ value in a future arterial blood gas measurement taken from the same patient. We used an existing opportunistic dataset of unselected ABG result pairs from hospitalised patients, described in detail previously.^17^ We quantified the median absolute error above baseline (MAE) in *P*_*a*_*O*_2_ to quantify predictive validity, with lower error values indicating better performance (Figure 1c, Supplementary Figure 2). Across a range of maximum cut-off values for *S*_*a*_*O*_2_, the lowest MAE value was obtained at 94% (Figure 1a; *S/F* MAE = 4.41 kPa (IQR: 2.74-6.63 kPa); *S/F*_94_ MAE = 3.32 kPa (IQR: 1.87-5.26 kPa), p(MWU) = 3.7 *×* 10^−18^).

**Figure 2:**
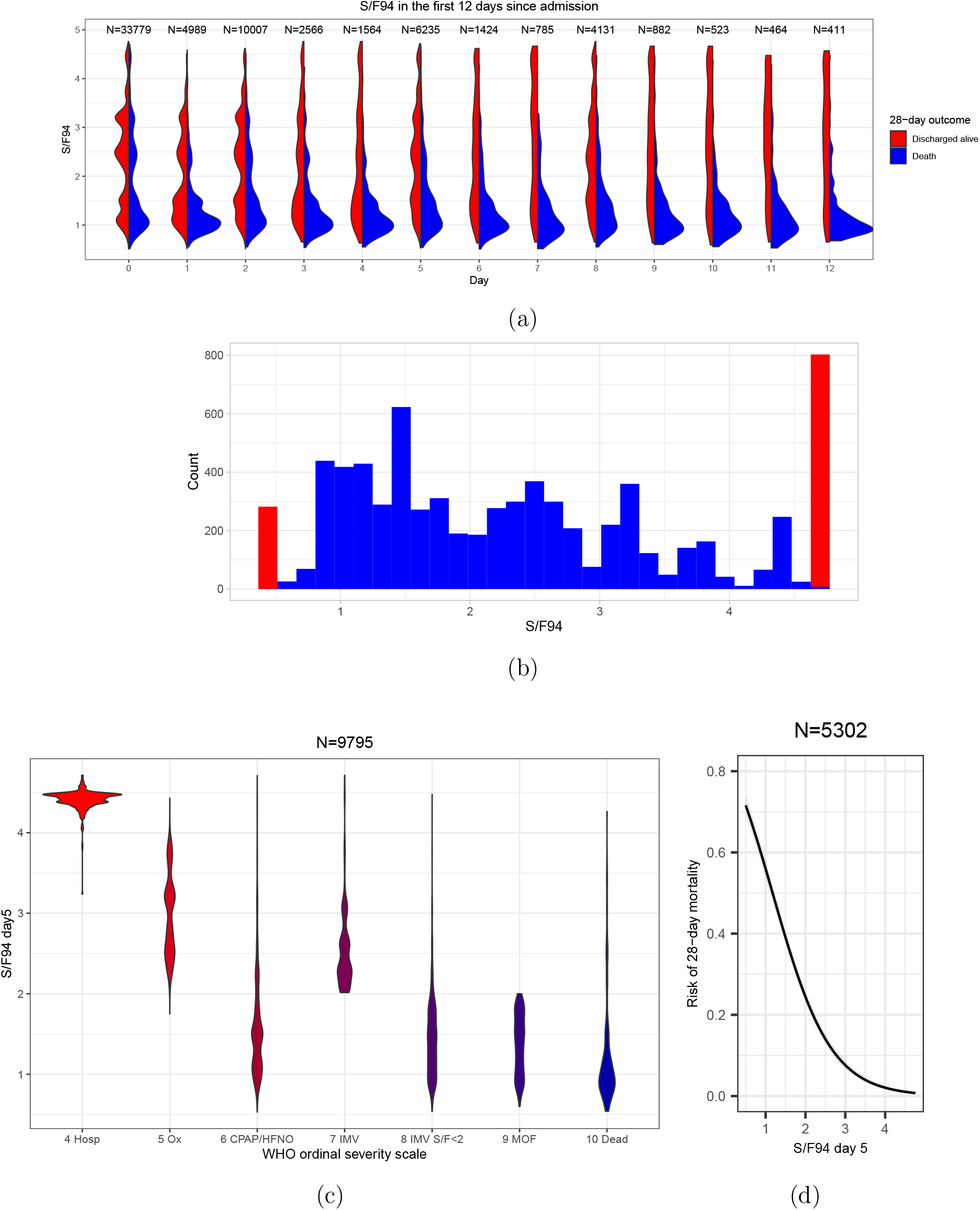
Evaluation of *S/F*_94_ in observational data. (a) Smoothed distributions of *S/F*_94_ values in survivors and non-survivors during the first 12 days of the study (restricted to 39,765 patients aged 20 − 75, oxygen therapy within 3 days). (b) Histogram showing distribution of *S/F*_94_ values on day 5 as used for subsequent analyses (in blue) Patients discharged home before day 5 are assigned the maximum value (4.78), and patients who died before day5 are assigned to an arbitrary minumum of 0.5 (in red). (c) Distribution of *S/F*_94_ values day 5 compared with WHO ordinal scale^2^ value at the same time point, in patients who met our inclusion criteria (aged 20 − 75, oxygen therapy within 3 days). For some patients who died on day 5, *S/F*_94_ values were available. For those with missing *S/F*_94_ values who died, an *S/F*_94_ of 0.5 was used to reflect poor pulmonary oxygenation function. Hosp = hospitalised, no oxygen support; Ox = Hospitalised, oxygen by mask or nasal prongs; CPAP/HFNO = Hospitalised, oxygen by continuous positive airway pressure; high-flow nasal oxygen or non-invasive ventilation; IMV = Intubation and mechanical ventilation; IMV *S/F<* 2 = Mechanical ventilation; *S/F <* 2 or vasopressors; MOF = Multi-organ failure & mechanical ventilation & *S/F <* 2 & ECMO or renal replacement therapy. (d) Logistic regression analysis with 95% confidence interval, using both *S/F*_94_ on day 0 and *S/F*_94_ on day 5 as covariates, showing a clear association between mortality at 28 days and *S/F*_94_ value on day 5.

### Evaluation in ISARIC4C data

39,765 cases in the ISARIC4C study had *S*_*a*_*O*_2_, *F*_*I*_*O*_2_ and clinical data available for analysis and met inclusion criteria (See Methods). Mortality in this population was 20.8% (Table 1). Since measurement of *S/F*_94_ was not protocolised in ISARIC4C, measurements were obtained for patients for whom *S*_*a*_*O*_2_ happened to be *<* 0.94 or who were breathing room air (*F*_*I*_*O*_2_ = 0.21), therefore meeting the *S/F*_94_ definition. The conceptual advantage of *S/F*_94_ over *S/F* is that it offers a closer relationship to the pathophysiological process of interest. This is not expected to be apparent in the distribution of values observed, but rather in the sensitive detection of a real therapeutic effect. For this reason, and because of the risk of selection bias (see Methods), we did not undertake a direct comparison of patients meeting the criteria for *S/F*_94_ measurement, against patients who do not. Instead, we evaluated *S/F*_94_ against other commonly used outcome measures.

**Table 1:**
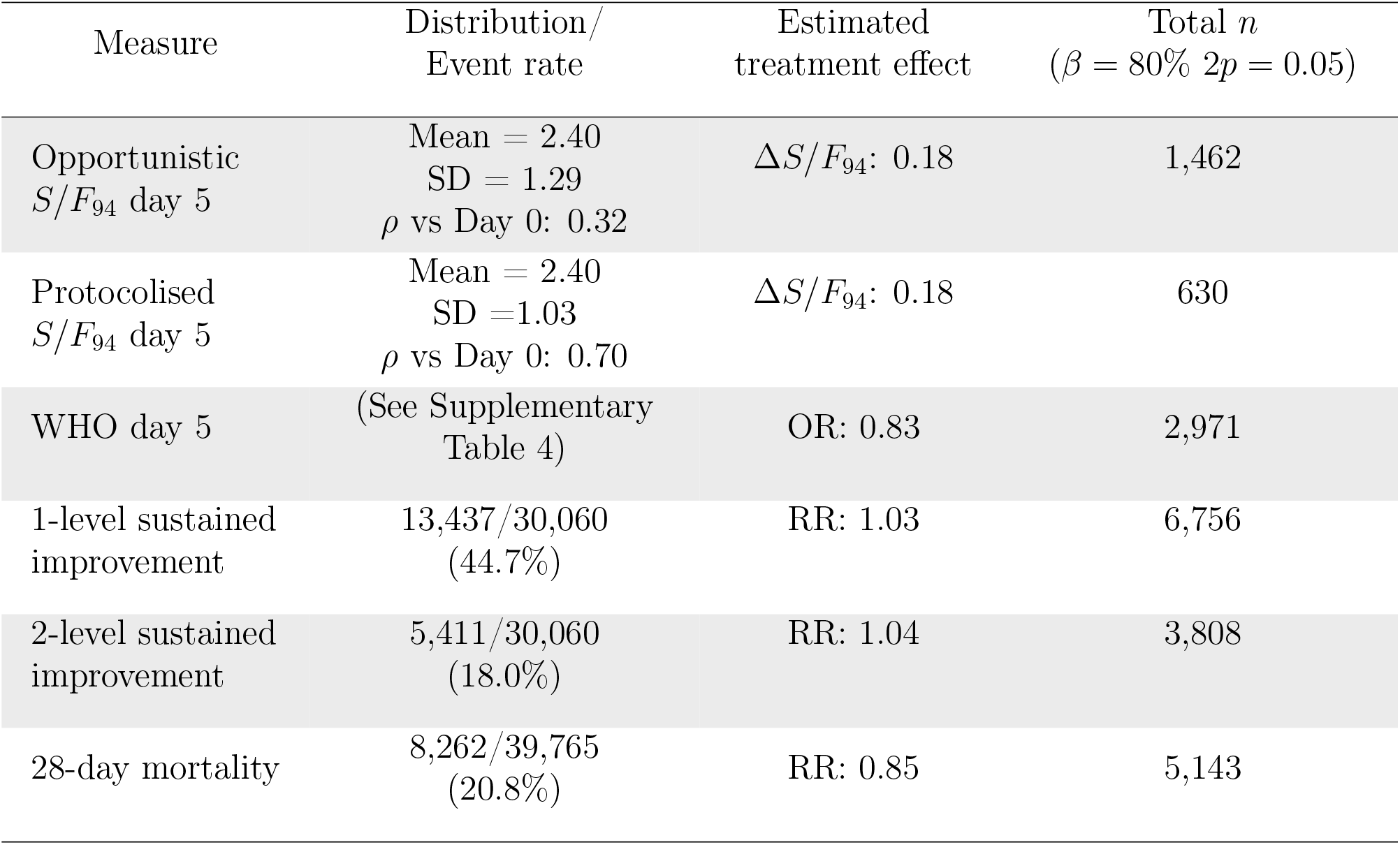
Comparison of outcome measures among 39,765 hospitalised patients aged 20-75, who required supplemental oxygen in the first 3 days in hospital. The estimated treatment effect is for a 15% relative reduction in mortality. Sample size shows the total number of subjects needed in both arms using a 1:1 allocation. Protocolised *S/F*_94_ - hypothetical improvement in power using a protocolised measurement of *S/F*_94_. Δ*S/F*_94_ - change in *S/F*_94_ associated with a 15% reduction in mortality. RR - risk ratio. OR - proportional odds ratio.

Within the ISARIC4C dataset, S/F values were available for the largest numbers of patients on days 0, 2, 5 and 8 from study enrolment. Among patients who remained in hospital, the distribution of *S/F*_94_ values moves over the first 5 days from study enrolment towards a bimodal pattern with high values in survivors, and low values in non-survivors (Figure 2a). We therefore chose day 5 as the primary timepoint for comparison.

An intermediate clinical outcome should have a strong association with a definitive outcome. Using 28-day mortality as the definitive outcome, and including *S/F*_94_ values on both day 0 and day 5 as covariates in a linear regression model, we found a strong inverse association between *S/F*_94_ on day 5 and mortality: an increased risk of mortality at day 28 is associated with a lower value of *S/F*_94_ on day 5 (Figure 2d). The OR for 28-day mortality is 0.25 (95% confidence interval 0.23-0.28), meaning that for a 1 unit increase in *S/F*_94_ on day 5, the odds of 28-day mortality decrease by 75%.

We also compared *S/F*_94_ with a widely-used intermediate outcome, the WHO scale. Since this scale records clinical decisions about therapy that are, in part, determined by the severity of hypoxic lung disease, a close relationship was expected with *S/F*_94_ (Figure 2c). The distributions were consistent between patients meeting the inclusion criteria (Figure 2c) and unselected patients (Supplementary Figure 5a).

### Sample size estimation

Using the observed relationships in ISARIC4C data for eligible patients (see Methods), we quantified effect sizes associated with a 15% relative risk reduction in mortality for each of the following measures: *S/F*_94_ at 5 and 8 days after study enrolment, the WHO ordinal scale at 5 and 8 days after study enrolment, the proportion of patients who reached a sustained 1 or 2-level improvement on the WHO ordinal scale, and a gold standard, 28-day mortality. We chose a 15% relative risk reduction in mortality based on previous power calculations for the RECOVERY trial. We then estimated the sample sizes required to detect these effects with 80% power at 2*p* = 0.05.

Some examples of sample size calculations using different inclusion criteria can be found in the supplemental information. We created an online tool, using synthetic data with similar characteristics to the ISARIC4C data (See methods), to enable users to test any combination of inclusion criteria (age, frailty score and type of respiratory support) and outcome assessment timepoint: https://isaric4c.net/endpoints.

For a 15% relative reduction in mortality, the required sample size was smallest for *S/F*_94_ on day 5, needing 731 patients in each arm (1,462 in total, Table 1). The number of subjects required for *S/F*_94_ on day 8 was higher, with 1,322 subjects in each arm. For the WHO ordinal scale, 1,486 participants would be required in each arm on day 5, or 1,161 on day 8 to detect this mortality reduction. Sample size was larger when 1-level sustained improvement was used as the outcome variable, with 3,378 patients in each arm, and 1,904 subjects in each arm when using 2-level sustained improvement.

#### Estimated improvement with protocolised measurement of *S/F*_94_

We have developed a protocol for measurement of *S/F*_94_ (Appendix: Protocol). Opportunistic measurements of *S/F*_94_ are likely to be less precise than protocolised measurements of *S/F*_94_, and hence to underestimate the relationship with the outcome variable, mortality.^18^ As a consequence, we expect a smaller number of participants will be required to detect a treatment effect at a given power when using protocolised *S/F*_94_ measurements as an outcome. We sought to estimate the magnitude of this improvement. Protocolising measurements is likely to substantially improve the accuracy of measurements of oxygenation function, firstly by ensuring that an oxygen delivery mode is used for which *F*_*I*_*O*_2_ can be accurately quantified (e.g. Venturi systems), and secondly by ensuring that measurements are taken at steady state. Protocolised measurement also permits inclusion of all patients, since *F*_*I*_*O*_2_ is decreased until *S*_*a*_*O*_2_ *<* 0.94, to a minimum of *F*_*I*_*O*_2_ = 0.21. A description of the estimation of effect size for the protocolised *S/F*_94_ measurement can be found in the supplemental methods. Based on this effect size estimate, sample size for a protocolised measurement of day 5 *S/F*_94_ would be around 630 subjects in total (Figure 3).

**Figure 3:**
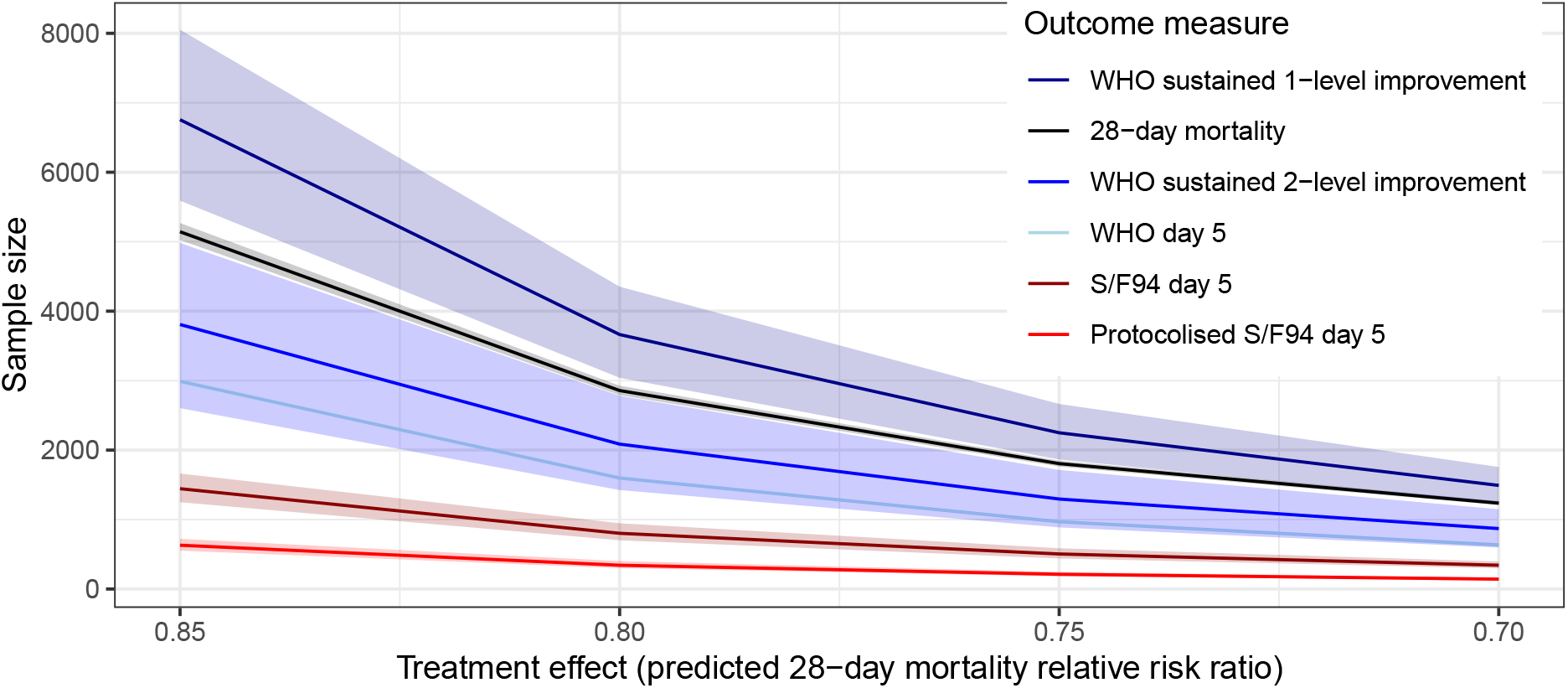
Comparison of the number of patients needed for the different outcome measures, using treatment effects between 0.85 and 0.70. The bottom line shows predicted sample size required when using a protocolised *S/F*_94_ measurement, rather than an opportunistic measurement.

## Discussion

We found that *S/F*_94_ performs well as a noninvasive measure of lung oxygenation function. *S*_*a*_*O*_2_ *<* 0.94 is an pragmatic cut-off value in a safe range, excluding the majority of obviously misleading values caused by the ceiling effect, and optimising predictive validity (Figure 1). *S/F*_94_ fulfils the criteria of an intermediate outcome: a continuous outcome measure that is closely related to mortality and can be modified by therapy.^3^ In a clinical trial setting, where both *S*_*a*_*O*_2_ and *F*_*I*_*O*_2_ measurement can be better protocolised, both the variance of *S/F*_94_, and the strength of the relationship to mortality are expected to improve. Comparing both the WHO ordinal scale and *S/F*_94_ to the definitive outcome of mortality at day 28, we found that the same predicted treatment effect can be detected with fewer patients using *S/F*_94_, even when measurement is not protocolised.

Our analyses may underestimate the statistical power of mortality, since time-to-event analyses would be used in most circumstances to maximise statistical power. Due to the large proportion of missing data after day 10, it was not possible to carry out survival modelling in our data. Ideally, we would have performed a mediation analysis with treatment effect, to determine the extent to which the treatment effect on mortality is explained by the intermediate endpoint *S/F*_94_. However, since there is no *S/F*_94_ data available from clinical studies showing treatment effect, it is not possible to perform this analysis.

*S*_*a*_*O*_2_ and *F*_*I*_*O*_2_ are both subject to measurement error, particularly in opportunistic data. Estimating *F*_*I*_*O*_2_ for patients receiving supplemental oxygen via nasal cannula or simple (Hudson) masks is inaccurate, because the *F*_*I*_*O*_2_ is profoundly affected by respiratory rate, tidal volume and inspiratory flow rate. The position of a patient on the ordinal WHO scale is influenced by both availability of resources and the decision by the patient and the clinician to not to escalate the level of care or provide organ support. This may explain the wide range of *S/F*_94_ values for patients at the same position on the WHO scale (Figure 2c).

There are multiple advantages of using *S/F*_94_ as an intermediate outcome measure in a phase II clinical trial in hospitalised patients. It is an easy, non-invasive measurement, using near-ubiquitous monitoring equipment. In contrast, daily *P*_*a*_*O*_2_ measurements (from an arterial blood sample) are time-consuming, require highly-skilled staff, and are burdensome for patients unless an indwelling arterial catheter is present (unusual outside of critical care areas).

Of the pragmatic endpoints available from routinely collected data, the WHO ordinal scale is the best-performing endpoint. In studies where clinical observations can be obtained, *S/F*_94_ is a robust measure of lung oxygenation function, and is the best measure to optimise statistical power for comparisons. *S/F*_94_ is comparable to the P/F ratio as a measure of lung oxygenation, and superior to *S*_*a*_*O*_2_/*F*_*I*_*O*_2_ ratio. Where protocolised measurements can be obtained, further improvements in statistical power are expected. *S/F*_94_ is a powerful and robust intermediate endpoint for clinical studies of COVID-19 and other causes of acute lung injury.

## Methods

### Relationship to gold standard (*P/F* ratio)

We evaluated the relationship between *S/F* and *P/F* in two datasets: a synthetic dataset of 1,529,176 predictions covering a wide range of possible physiological variation, generated by a mathematical model of oxygen delivery written in Python (available at https://github.com/baillielab/oxygen_delivery) and reported previously,^17^ and 72,457 unselected arterial blood gas results from a critically ill population.^17^ Taking *P/F* to be the gold standard, we evaluated *S/F* at different thresholds in both synthetic and real data.

### Predictive validity

We considered the predictive validity of *S/F* and *S/F*_94_ compared to *P/F* and two other measures of oxygenation function: the Alveolar-arterial difference (A-a), and effective shunt fraction (ES).^17^

Predictive validity quantifies the extent to which a clinical measurement predicts an unseen event. The aim is not to optimise prediction, but to test the extent to which a measurement is describing a real feature of the patient’s illness.^19^ In this case, we contend that a measure that accurately describes lung oxygenation function will accurately predict *P*_*a*_*O*_2_ after a change is made to *F*_*I*_*O*_2_. Using the same opportunistic dataset as in our previous study,^17^ we used this approach to assess the validity of *S/F* and *S/F*_94_.

Briefly, in pairs of arterial blood gas (ABG) results taken from the same patient <3h apart, in which *F*_*I*_*O*_2_ was decreased in the later sample (indicating weaning, and hence clinical stability), we used various measures of oxygenation (A-a, *P/F*, ES, *S/F*) in the first ABG to predict the *P*_*a*_*O*_2_ in the second sample. Predictive validity is quantified by the median absolute error (difference from the real value). A baseline value, showing the difference between ABG results for matched pairs in which *F*_*I*_*O*_2_ did not change, is provided to contextualise the MAE results by providing a reasonable minimum error value as a baseline. Results are presented as difference in MAE from this baseline (MAE). Mann-Whitney U-test (MWU) was used for comparison of MAE difference from baseline.

### Evaluation in ISARIC4C data

#### Inclusion criteria

All subjects were part of the ISARIC Coronavirus Clinical Characterisation Consortium (ISARIC4C) WHO Clinical Characterisation Protocol UK (CCPUK), a study in England, Wales, and Scotland prospectively collecting data from patients hospitalised with SARS-CoV-2 infection since the start of the pandemic.

In order to focus our assessment on the subset of patients with hypoxaemic respiratory failure that is potentially modifiable by anti-inflammatory treatment, we repeated all analyses in subjects aged 20-75 who required supplementary oxygen therapy within 3 days of hospital admission, subjects aged 20-75 that were oxygen dependent on the day of admission, and subjects aged 20-75 without criteria for oxygen dependency. All included patients had *S*_*a*_*O*_2_ and *F*_*I*_*O*_2_ data available.

#### Estimation of *S/F*_94_ in observational data

The *S/F* ratio was calculated by dividing *S*_*a*_*O*_2_ by *F*_*I*_*O*_2_ (with both as fractions taking values between 0 and 1). For this evaluation, *S/F*_94_ was defined as an opportunistic measurement in which *S*_*a*_*O*_2_ ≤ 0.94, or the patient was receiving no supplementary oxygen (*F*_*I*_*O*_2_ = 0.21).

Importantly, the retrospectively-defined subgroup of patients meeting the *S/F*_94_ criteria is not representative of all patients since there was an excess of patients who were not receiving respiratory support, with slight excess mortality, in the *S/F*_94_ group (Supplementary Table 1). This indicates at least two mechanisms of selection bias, acting in opposite directions, and precluding a direct comparison. Firstly, patients who have high blood oxygen levels on relatively little supplementary oxygen are excluded from the *S/F*_94_ group; by definition these patients have relatively mild disease. Secondly, the group in whom *S/F*_94_ could be measured includes patients who receive supplemental oxygen, and fail to reach adequate *S*_*a*_*O*_2_ values, but are not escalated to a higher level of respiratory support; this is a frail and multimorbid population with very severe disease.

*S/F*_94_ was calculated at baseline (day 0) and on day 5 and day 8 from study enrolment. There is expected to be differential missingness between *S/F*_94_ and mortality: *S*_*a*_*O*_2_ and *F*_*I*_*O*_2_ data are only available for a proportion of cases, whereas outcome data is well-recorded. Patients who died or were discharged on given day and had a missing value for *S/F*_94_ were assigned values 0.5 (severe oxygenation defect) and 4.76 (perfect oxygenation), respectively. However, death/discharge was more likely to be recorded than *S/F*_94_, and this could introduce bias into our analysis. We addressed this by estimating the proportion of patients for whom *S/F*_94_ measurements were available and who would be expected to die/be discharged at a given point in time. We then resampled those who died/discharged according to these proportions. For example, if on day 5 5% of patients had died, and 15% went home, the other 80 % was still hospitalised. We resampled from those who died/ were discharged alive, so that the non-missing values reflected the same proportion (5% dead with *S/F*_94_ set to 0.5, 15% discharged alive with *S/F*_94_ set to 4.76 and the other 80 % still hospitalised with available *S/F*_94_ values.

#### Association between *S/F*_94_ and 28-day mortality

Two key assumptions underlie the use of *S/F*_94_ as an intermediate endpoint. Firstly, that pulmonary oxygenation function lies on the causal pathway to death in COVID-19, and secondly, that *S/F*_94_ accurately reflects pulmonary oxygenation function. If either of these assumptions are violated, then a strong relationship between *S/F*_94_ and subsequent mortality would not be expected.

To evaluate this association, a logistic regression model was developed with 28-day mortality as the dependent variable and *S/F*_94_ measured on day 0 and day 5 as two separate covariates. Given the strong relationship between *S/F*_94_ on day 0 and *S/F*_94_ on days further in the disease trajectory, we included both *S/F*_94_ on day 0 and day 5 as covariates. Linear dependence of log-odds on *S/F*_94_ measured on day 0 and day 5 was assessed both by visual inspection and with model selection criteria including the Bayesian Information Criterion (BIC). Nonlinearities were evaluated using a restricted splines model. Finally, predicted models were made to assess the absolute change in risk of mortality with a change in *S/F*_94_.

#### Sample size calculations

We compared the sample sizes required for a range of different outcomes measures (*S/F*_94_, WHO ordinal scale, sustained improvement at day 28 and 28-day mortality). For the intermediate endpoints, we estimated the treatment effect associated with a 15% relative reduction in mortality. Below we give brief descriptions of the effect size calculations for the different outcome measures. All calculations assumed a 1:1 allocation of participants between treatment and control groups and are based on having 80% power at p= 0.05 (two-sided) to detect the stated treatment effect. Details on effect size estimation can be found in the supplementary material.

#### Quantifying uncertainty

Errors around the point estimates shown in Table 1 are shown in Figure 3 for a range of effect sizes. The upper and lower limits of the 95% confidence interval calculated for the effect size were used to bootstrap 95% confidence intervals for sample size. This 95% confidence interval does not capture uncertainty around validity of the modelled relationship between outcome and effect. The sample size and 95% confidence interval when the outcome measure is 1- or 2-level sustained improvement on the WHO scale are noticeably larger than for the other outcome measures, in particular *S/F*_94_. There are several reasons for this. The first is that the magnitude of effect size compared to variance of outcome measure is much smaller when 1- or 2-level sustained improvement on the WHO scale is the outcome measure compared to when *S/F*_94_ is the outcome measure. The second reason is that *S/F*_94_ is a continuous variable, whereas 1- or 2-level sustained improvement on the WHO scale is a Bernoulli variable whose mean is the proportion of people who had a 1 or 2-level sustained improvement on the WHO scale, respectively. The sample size calculation with *S/F*_94_ as outcome measure relies on a two-sample t-test for testing the hypothesis that the means of two normal distributions with the same variance are equal. In the case of 1- or 2-level sustained improvement on the WHO scale, the procedure is similar except the variances of the two distributions are not equal. The mean of a large number of independent Bernoulli variables with mean *μ* is approximately normally distributed with mean *μ* and variance *μ*(1 − *μ*). Therefore, two sets of Bernoulli distributed variables with different means also have different variances. If we use *μ*_1_ to denote the mean for the control group, and *μ*_2_ to denote the mean for the treatment group, then while *μ*_1_ *< μ*_2_ ≤ 0.5, the variance of the sample mean of the outcome measure increases as the effect size *μ*_2_ − *μ*_1_ increases. Thus while these conditions hold, and with all else equal, this has the consequence that an outcome measure that is Bernoulli distributed will require larger sample sizes than an outcome measure that has a continuous distribution.

#### Continuous variables vs (*S/F*_94_)

We fit a logistic regression with mortality at day 28 as the independent variable, and *S/F*_94_ on day 0 (baseline) and day 5 (or day 8) as predictors. Age and sex were also included in the model as they are strong predictors of mortality. We used this to calculate the predicted probability of mortality, and the change in *S/F*_94_ associated with a relative reduction in predicted mortality of 15%, for each subject. Finally, we took the mean to find the average change in day 5 *S/F*_94_ that is associated with a 15% reduction in mortality across the sample. This is the target treatment effect in the clinical trial. We calculated the sample size required to see this treatment effect with a given level of power using a two sample t-test with ANCOVA correction for the correlation between *S/F*_94_ on day 0 and day 5.^20^

#### Ordinal variables (WHO scale)

Values for the WHO ordinal scale were derived using information about oxygen support and mortality. Possible values in hospitalised patients range between 4 and 10.^2^

#### WHO scale - absolute value

Sample size calculations for this outcome are based on the proportional odds model. In order to estimate the odds ratio equivalent to a 15% relative reduction in mortality, a proportional odds model was used.^21^ The dependent variable was WHO ordinal scale on day 5 or 8, with age and sex as predictor variables.

#### WHO scale - sustained improvement

We calculated the number of patients who had a sustained 1- or 2-level improvement in the WHO scale. To be considered sustained, an improvement had to be maintained until discharge or until day 28. We calculated sample size for this outcome using a two-sample test for proportions with a continuity correction.^22^ Only patients who had WHO ordinal scale values on at least two separate days were included in this analysis.

We used a logistic regression model to analyse the relationship between mortality and the predictors sustained improvement, age and sex. We then used this model to estimate the difference in proportion of people who had a sustained improvement on the WHO ordinal scale that is associated with a 15% reduction in risk of mortality.

#### Mortality

In order to compare these alternative outcome measures with a gold standard (mortality), we calculated the number of participants needed if 28-day mortality was the outcome measure, using a two-sample test for proportions with continuity correction.

## Supporting information

SF94 measurement protocol

## Data Availability

The dataset used and analysed in this study is not available, but a synthetically generated dataset, containing the same key properties as the original dataset is available for sample size calculations on https://isaric4c.net/endpoints

## Declaration

### Ethics approval

Ethical approval was given by the South Central-Oxford C Research Ethics Committee in England (13/SC/0149), the Scotland A Research Ethics Committee (20/SS/0028), and the WHO Ethics Review Committee (RPC571 and RPC572, April 2013).

### Authors’ contributions

JKB and PH concieved the study. JKB, MGS and PJMO acquired funding. JKB, PWH, FM, NY, JD, ADB, JM, JSN-V-T, PWH, and MGS designed the analysis. EMH, RG, ES, ABD, DH, NS and NL provided guidance on methodology and interpretation. MCS, SK, ABB, NS and JKB did the formal analysis. JSB and SK created the website. EH, ABD, GHG, NL, NS and JKB supervised the work. MCS, SK and JKB wrote the original draft of the manuscript. All authors reviewed and gave feedback on the manuscript. All authors read and approved the final manuscript.

## Acknowledgements

This work uses data provided by patients and collected by the NHS as part of their care and support. We are extremely grateful for the front-line NHS clinical and research staff and volunteer medical students who collected this data in challenging circumstances, and the generosity of the participants and their families for their individual contributions in these difficult times. We also acknowledge the support of Jeremy J Farrar (Wellcome Trust) and Nahoko Shindo (WHO). For the purpose of open access, the author has applied a CC BY public copyright licence to any Author Accepted Manuscript version arising from this submission.

