## Supplementary material for "Evaluation of *S*/*F*_94_ as a proxy for COVID-19 severity": SF94 measurement protocol

### Clinical protocol for measurement of lung oxygenation (S/F94)

#### The S/F94 clinical outcome measure

This is a protocol for measurement of lung oxygenation function as an outcome measure for a clinical study. S/F ratio is a simple correction for the measured $S_pO_2$ to take into account how much oxygen the patient is receiving ($F_IO_2$). It is simply calculated as $S_pO_2$ divided by $F_IO_2$. But if the patient's $S_pO_2$ is high, S/F stops being an accurate reflection of lung oxygenation function. This is because, above 94%, the $S_pO_2$ value can't rise much, no matter how much oxygen the patient is receiving (sec. 2). S/F94 fixes this by turning down the oxygen until the patient's $S_pO_2$ is below 94%.

**S/F94** is the S/F ratio measured when the patient's $S_pO_2$ is below 94% on any oxygen therapy, or at any $S_pO_2$ at when they are breathing air.

#### How to measure

Essential conditions:

1. Oxygen therapy is giving a **measurable percentage of oxygen** (sec. 1.1).
2. Patient has been **resting, not talking, on the same oxygen therapy or at least 5 minutes** (sec. 1.4).
3. Either **$S_pO_2$ is less than 94%**, or the patient is breathing air.

If these conditions are met:

4. Complete the CRF to record $S_pO_2$, the mode of oxygen delivery, and the percentage of oxygen being delivered ($F_IO_2$).
5. Revert to original oxygen therapy once the measurement is complete.

#### Exclusions

This protocol should not be used in patients undergoing treatment for acute brain injury or post-cardiac arrest.

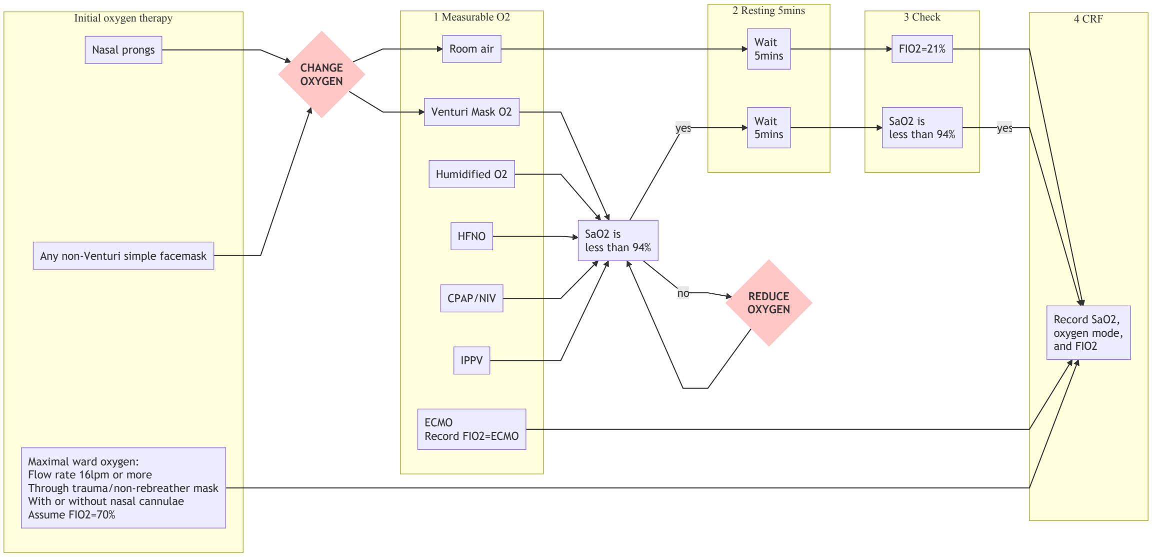


Figure 1: Flowchart for measurement of S/F94

### 1 Additional information and procedures

#### 1.1 Modes of oxygen delivery that give a measurable percentage of oxygen

If the patient is not receiving a measurable percentage of oxygen, change to a different mode of oxygen therapy: sec. 1.2.

| Acceptable oxygen modes | Description | $F_IO_2$ to record |
|---|---|---|
| Room air | $S_pO_2$ on air is always acceptable if the patient has been breathing air for at least 5 minutes. | 21% |
| Venturi Mask | Venturi masks have a changeable attachment on the oxygen inflow stating an oxygen percentage. | 24%, 28%, 31%, 35%, 40%, 60% |
| Trauma mask ± nasal prongs | A trauma mask (one with a non-rebreathing bag) is often used, sometimes together with nasal prongs, to provide maximal oxygenation for a ward patient. If this can't be reduced safely, $F_IO_2$ should be recorded at 70% | 70% |
| HFNO | High-flow nasal oxygen | 21-100% |
| Humidified $O_2$ | A high-flow humidified system providing a specified oxygen percentage | 21-100% |
| CPAP/NIV | Any kind of continuous positive airway pressure or non-invasive respiratory support | 21-100% |
| IPPV | Invasive positive pressure ventilation through an endotracheal tube or tracheostomy | 21-100% |
| ECMO | Extra-corporeal membrane oxygenation | ECMO |

#### 1.2 Changing oxygen

If the patient is receiving oxygen by nasal prongs or any simple non-Venturi mask, change to an acceptable oxygen system as clinically appropriate. Some suggestions are below:

| Previous mode | $S_pO_2$ | Change to |
|---|---|---|
| Nasal prongs less than 4l/min | above 90% | Room air |
| Nasal prongs less than 4l/min | above 86% | Venturi mask 24% |
| Simple (Hudson) facemask less than 4l/min | above 90% | Room air |
| Simple (Hudson) facemask less than 4l/min | above 86% | Venturi mask 28% |
| Other masks with flow up to 15l/min | above 90% | Venturi mask 40% |
| Other masks with flow up to 15l/min | above 86% | Venturi mask 60% |
| Other masks with flow at or above 15l/min | below 86% | Do not reduce $O_2$. Record $F_IO_2$ as 70% |

#### 1.3 Reducing oxygen

If the patient remains comfortable, $S_aO_2$ values as low as 80%, particularly for short periods (less than 20mins), are not thought to be harmful. The patient should be watched continuously for signs of distress after each change to oxygen therapy. Cyanosis (blue colouration of the lips) is normal in patients with $S_aO_2$ values in the range expected for this measurement.

If $S_pO_2$ is above 94%, reduce the $F_IO_2$ and monitor $S_pO_2$ continuously for 5 minutes.

| Previous $F_IO_2$ | $S_pO_2$ | Change to $F_IO_2$ |
|---|---|---|
| 30% or less | above 94% | Room air |
| 40% or less | above 94% | 28-30% |
| 60% or less | above 94% | 40% |
| 80% or less | above 94% | 60% |
| 100% or less | above 94% | 80% |

If the patient becomes breathless, agitated or feels unwell after a change in oxygen therapy, immediately revert to the previous oxygen therapy.

#### 1.4 Resting

The patient should be supine in bed or in a chair. If they are in bed, the head of the bed should be tilted upwards at $\geq 30°$. The patient must not be talking or exercising during this time.

#### 1.5 Special considerations

##### 1.5.1 Delirium/dementia

If patients are agitated this measurement will be very difficult and so the requirement that the patient should be resting and not talking will have to be relaxed. As long as the other criteria are met (patient is receiving a measurable percentage of oxygen, and $S_pO_2$ is less than 94% or the patient is breathing air)

##### 1.5.2 Talking

Our experience is that many Covid patients are keen to talk when we first meet them. If they meet the 3 essential conditions (How to measure), then a $S_pO_2$ measurement can be taken immediately after you introduce yourself. Otherwise, explain to the patient that talking may alter their oxygen levels and so they must remain calm and silent for 5mins.

##### 1.5.3 Proning

Patients who are being managed prone (either awake proning or on IPPV) should have S/F94 measurements taken during the supine periods as part of standard care.

#### 1.6 CRF elements

- Delivery mode
- $S_aO_2$
- $F_IO_2$
- HFNO flow rate
- PEEP (CPAP/NIV/IPPV)
- RR (as an alternative endpoint)

### 2 Rationale

The $S_aO_2/F_IO_2$ ratio (S/F) is a continuous index of lung oxygenation function and can be calculated without an arterial blood gas sample. It correlates well with the most widely-used measure of oxygenation - $P_aO_2/F_IO_2$ ratio (P/F).[1] S/F under steady state conditions in humans can range from around 0.5 (severe oxygenation defect) to 4.8 (perfect oxygenation function).

A major limitation of S/F is the ceiling effect: at high $S_aO_2$ values, $S_aO_2$ ceases to be dependent on lung oxygenation function, because the blood is close to maximally-oxygenated. A healthy patient with perfect lungs breathing 21% oxygen with $S_aO_2$=0.99 would have S/F = 4.7; but the same patient breathing 100% oxygen would have S/F = 0.99.

The S/F94 measurement removes these erroneous values, providing better agreement with a gold standard measure of oxygenation function (P/F ratio, Figure 2).

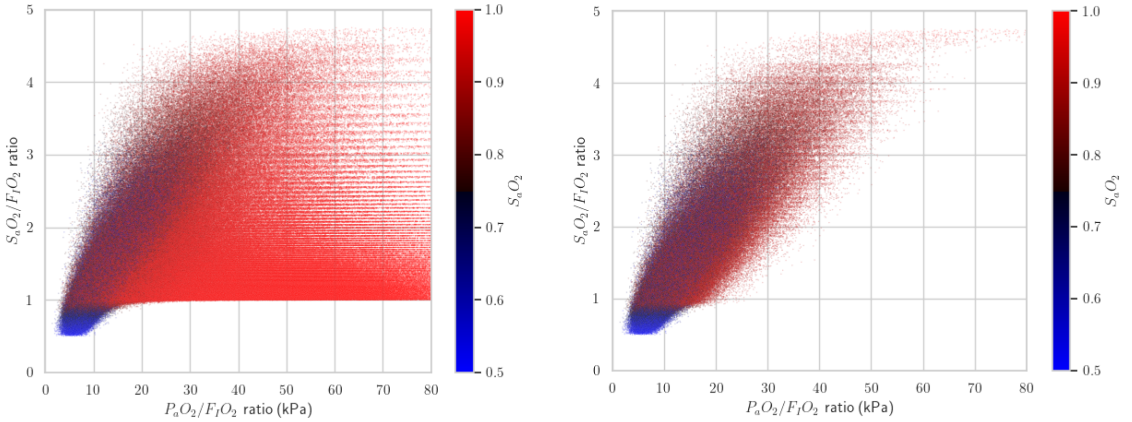


(a) All values (b) $S_aO_2 \leq 94\%$ or $F_IO_2 = 21\%$ only

Figure 2: Comparison of P/F and (a) S/F or (b) S/F94 in synthetic data generated using an computer model of gas exchange.[2] Points are coloured according the $S_aO_2$ as shown in the colour scale.

#### 2.1 Glossary

$S_aO_2$: *S*aturation of *a*rterial blood with oxygen ($O_2$).

$S_pO_2$: *S*aturation (measured by *p*ulse oximetry) of arterial blood with oxygen ($O_2$).

$F_IO_2$: *F*raction (*I*nspired) of oxygen ($O_2$).
