## Supplementary information for "Evaluation of *S*/*F*_94_ as a proxy for COVID-19 severity"

### 1 Supplementary information

#### 1.1 Physiology

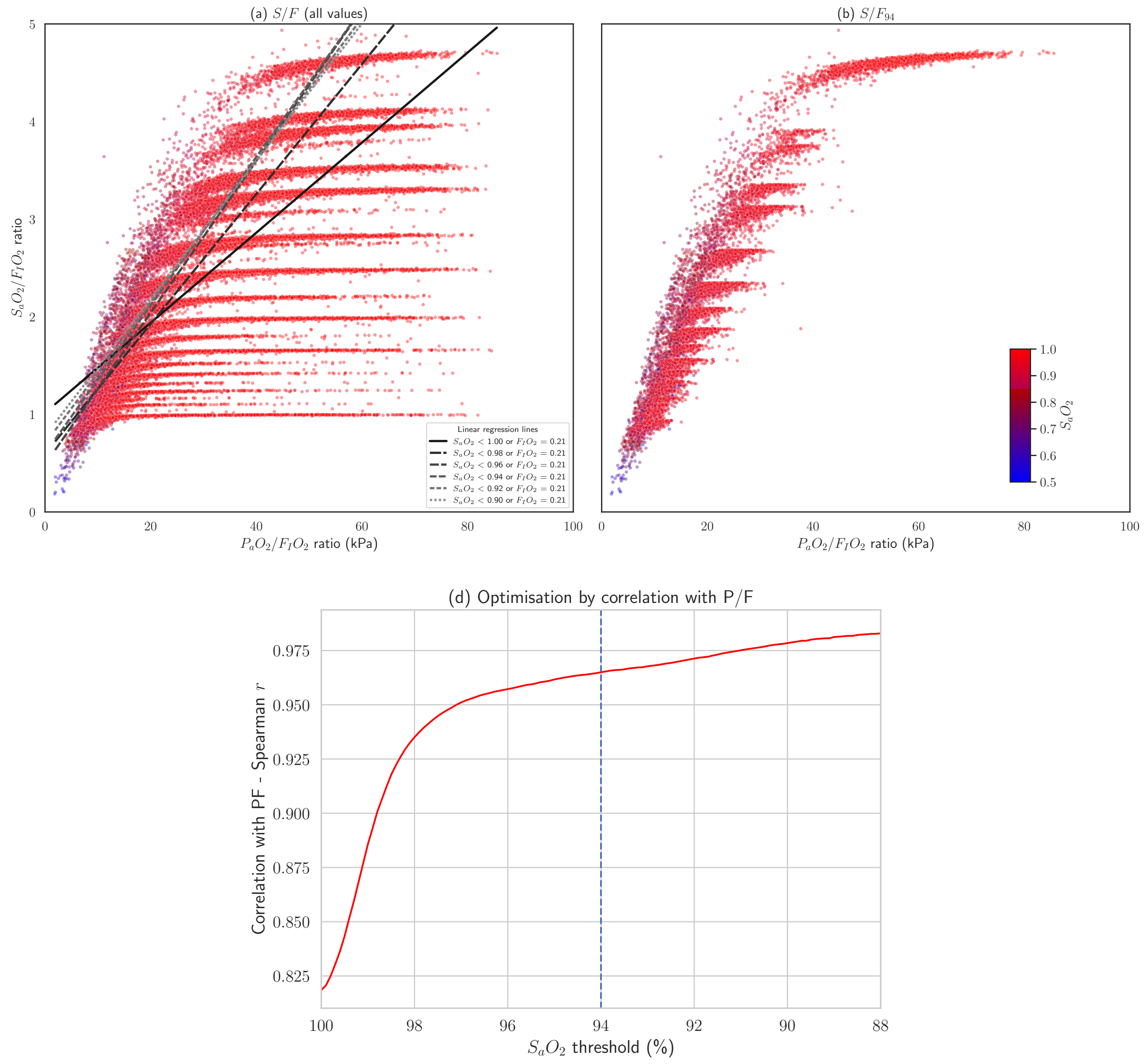


Supplementary Figure 1: (a,b) Scatter plots showing real blood gas results from critically ill patients. $S/F$ is plotted against a gold standard, $P/F$. Points are coloured according the $S_aO_2$ as shown in the colour scale. (a) including all values with no maximum $S_aO_2$. Linear regression lines are shown for $S/F$ against $P/F$ in using different maxima for $S_aO_2$; patients breathing air ($F_IO_2$=21%) were included in all regression analyses regardless of $S_aO_2$. (b) shows only values meeting the criteria for $S/F_{94}$: $S_aO_2 < 94\%$ or $F_IO_2 = 21\%$ (c) Optimisation of cut-off value for $S_aO_2$ using change in correlation coefficient (Spearman $r$) as the threshold for inclusion is lowered from $S_aO_2$< 100% to $S_aO_2$< 80%.

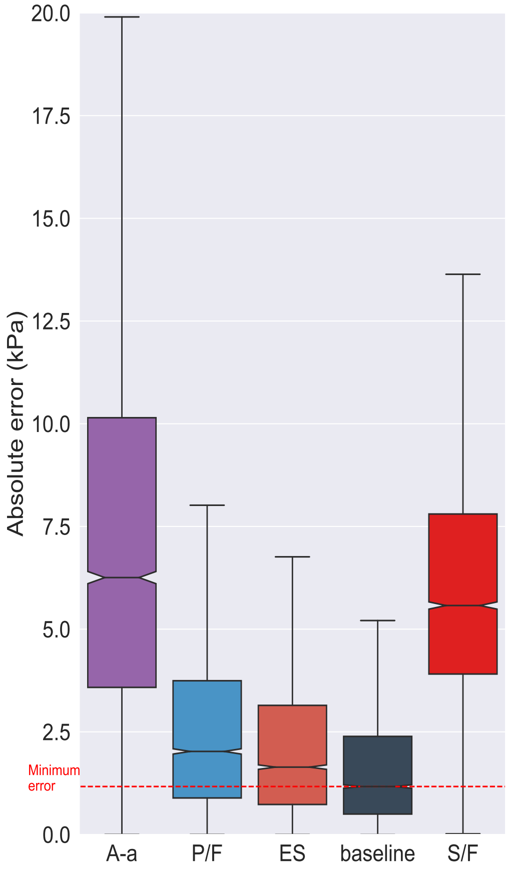


(a) Including all recordings

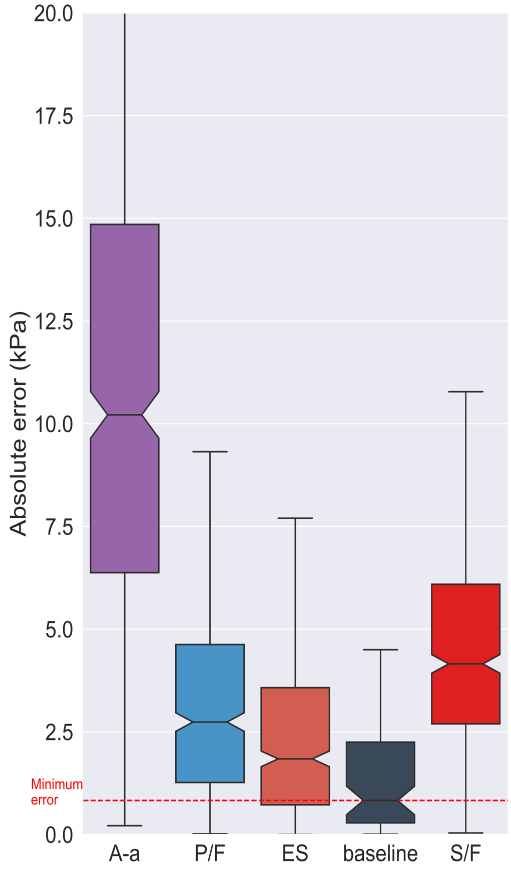


(b) Restricted to recordings where $S_aO_2 < 94\%$

Supplementary Figure 2: Predictive validity (ability to predict a future $P_aO_2$ after a change in $F_IO_2$; absolute error in kPa) of Alveolar- arterial (A-a), $P/F$ ratio ($P/F$), effective shunt fraction (ES), and $S/F$ ratio (S/F), compared to a baseline (the minimum achievable error, measured as the change in $P_aO_2$ between pairs of arterial blood gas measurements meeting the other selection criteria, with no change in $F_IO_2$). Lower values indicate superior validity. All values are included in (a); in (b) only values in which $S_aO_2 < 0.94$ in the first measurement.

#### 1.2 Evaluation in ISARIC data

##### 1.2.1 Respiratory Rate

There is large variation in respiratory rate, which is independent of $S/F_{94}$. As expected, there is a tendency of higher respiratory rates for patients with a lower $S/F_{94}$ (Supplementary Figure 3)

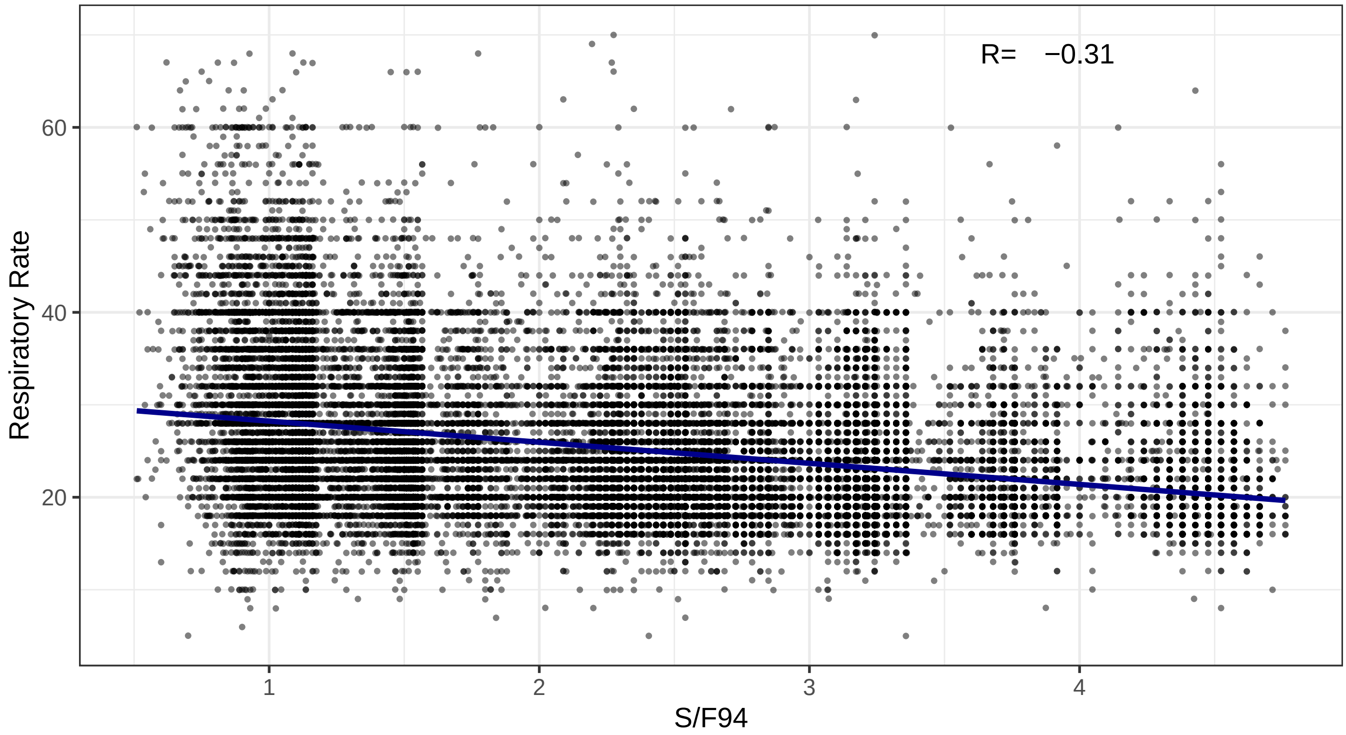


Supplementary Figure 3: Relationship between respiratory rate and the $S/F_{94}$ in 49,727 patients meeting inclusion criteria (age 20-75; oxygen therapy in first 3 days of admission).

###### 1.2.2 Association between mortality at day 28 and $S/F_{94}$

The relationship between $S/F_{94}$ on day 0 and mortality in the multivariable model appears counter-intuitive (Supplementary Figure 4a). A univariable regression model with mortality on day 28 as the outcome variable and $S/F_{94}$ on day 0 as the predictor, shows a relationship as expected: there is an increase in mortality risk with a decrease in $S/F_{94}$ on day 0 (Supplementary Figure 4b). Hence the reversed direction in the multivariable model is explained by the effect of change in $S/F_{94}$ between day 0 and day 5, suggesting that patients who are admitted to hospital with moderate oxygenation defects, and deteriorate rapidly, have a slightly increased risk of mortality.

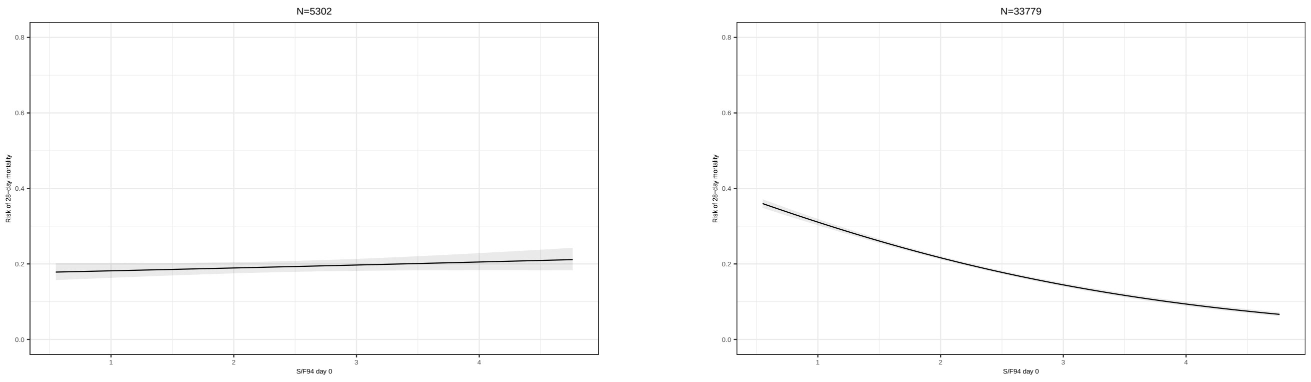


(a) Multivariate regression model

(b) Univariate regression model

Supplementary Figure 4: Results of regression model for $S/F_{94}$, using mortality at day 28 as the outcome, in (a) multivariable model including both day 0 and day 5 $S/F_{94}$ and (b) a univariate model with day 0 $S/F_{94}$ as the sole predictor variable. The univariate model shows a clear and expected association between mortality at day 28 and $S/F_{94}$ on day 0.

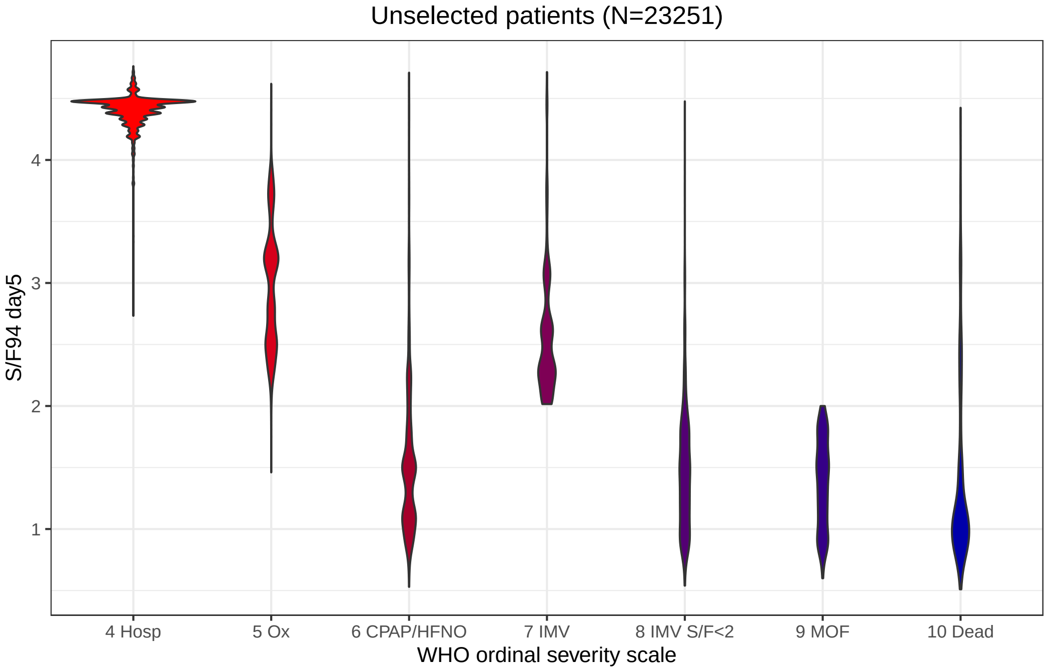


(a) $S/F_{94}$ on Day 5 by WHO ordinal scale.

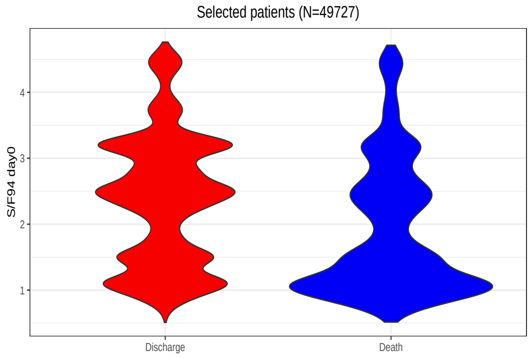


(b) $S/F_{94}$ on Day 0

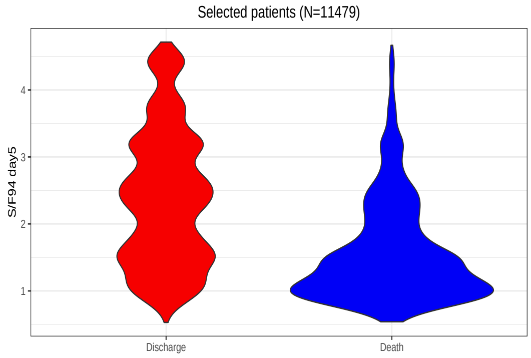


(c) $S/F_{94}$ on Day 5

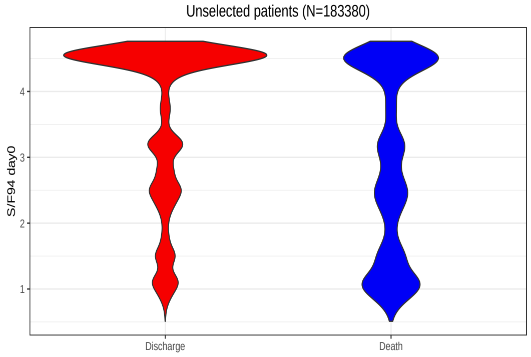


(d) $S/F_{94}$ on Day 0

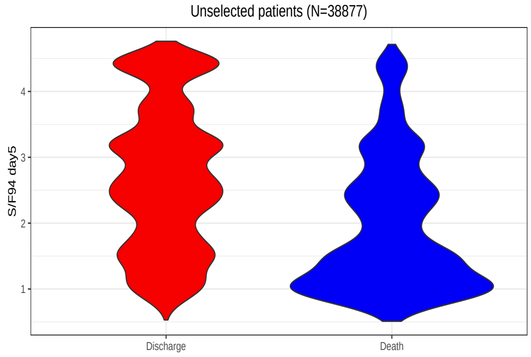


(e) $S/F_{94}$ on Day 5

Supplementary Figure 5: Relationship between (a) Distribution of $S/F_{94}$ values day 5 compared with WHO ordinal scale value at the same time point. For some patients who died on day 5, $S/F_{94}$ values were available. For those with missing $S/F_{94}$ values who died, an $S/F_{94}$ of 0.5 was used to reflect poor pulmonary oxygenation function. (b) $S/F_{94}$ on day 0 and 28-day outcome and (c) 28-day outcome and $S/F_{94}$ on day 5 among selected patients meeting inclusion criteria (age 20-75; oxygen therapy in first 3 days) (d) 28-day outcome and $S/F_{94}$ on day 0 and (e) 28-day outcome and $S/F_{94}$ on day 5 among unselected patients. Hosp = hospitalised, no oxygen support; Ox = Hospitalised, oxygen by mask or nasal prongs; CPAP/HFNO = Hospitalised, oxygen by continuous positive airway pressure; high-flow nasal oxygen or non-invasive ventilation; IMV = Intubation and mechanical ventilation; IMV $S/F \leq 2$ = Mechanical ventilation; $S/F \leq 2$ or vasopressors; MOF = Multi-organ failure & mechanical ventilation & $S/F_{94} < 2$ & ECMO or renal replacement therapy.

##### 1.2.3 Effect size estimation

This section details how effect sizes associated with a given treatment effect were calculated for different outcome measures.

###### 1.2.3.1 $S/F_{94}$ and sustained WHO ordinal scale improvement

We used a logistic model with mortality as the dependent variable, and separately estimated relationships with $S/F_{94}$ and WHO ordinal scale improvement,

$$\ln\left(\frac{p_i}{1-p_i}\right) = \beta_0 + \beta_1 y_i + \beta \cdot z_i. \tag{1.1}$$

Here, $p_i$ is the probability of day 28 mortality for individual $i$, $\beta_0$ and $\beta_1$ are scalar coefficients, $\beta$ is an $n$-dimensional vector of coefficients, $y_i$ is the outcome measure ($S/F_{94}$ or sustained WHO ordinal scale improvement encoded as a numeric variable taking values 0 or 1), and $z_i$ is an $n$-dimensional vector of covariates.

Let $\hat{\beta}_0, \hat{\beta}_1, \hat{\beta}$ be consistent estimators for $\beta_0, \beta_1, \beta$ respectively. $\hat{p}_i$ is defined as the corresponding predicted probability of day 28 mortality for patient $i$,

$$\ln\left(\frac{\hat{p}_i}{1-\hat{p}_i}\right) = \hat{\beta}_0 + \hat{\beta}_1 y_i + \hat{\beta} \cdot z_i. \tag{1.2}$$

We consider an individual-specific treatment effect given by

$$p_i \to f(p_i) \tag{1.3}$$

for some function f taking values in $[0, 1]$, that may in principle depend on all covariates. The changes $\Delta_i, \hat{\Delta}_i$ in the outcome measure associated with this treatment effect for individual $i$ keeping all other covariates constant in 1.1 and 1.2 satisfy

$$\ln\left(\frac{f(p_i)}{1-f(p_i)}\right) = \beta_0 + \beta_1 (y_i + \Delta_i) + \beta \cdot z_i. \tag{1.4}$$

$$\ln\left(\frac{f(\hat{p}_i)}{1-f(\hat{p}_i)}\right) = \hat{\beta}_0 + \hat{\beta}_1 (y_i + \hat{\Delta}_i) + \hat{\beta} \cdot z_i. \tag{1.5}$$

Our estimator for the expected change in $y_i$ associated with the treatment effect $f$ is obtained by subtracting 1.2 from 1.5, dividing by $\hat{\beta}_1$ and averaging over the sample,

$$\hat{\mu}_\Delta := \frac{1}{\hat{\beta}_1 I} \sum_{i=1}^{I} \ln\left(\frac{f(\hat{p}_i)(1-\hat{p}_i)}{\hat{p}_i(1-f(\hat{p}_i))}\right). \tag{1.6}$$

This is a consistent estimator for $\mu_\Delta = \mathbb{E}[\Delta_i]$, which follows from the fact that $\hat{\beta}_0, \hat{\beta}_1, \hat{\beta}$ are consistent estimators for $\beta_0, \beta_1, \beta$ and assuming $\{\Delta_i\}_{i=1}^{I}$ is mean ergodic.

In the results presented in this paper, a treatment effect that is a relative risk reduction of e.g. 15% means that $f$ is defined by

$$f(x) = 0.85x. \tag{1.7}$$

That is, the patient's predicted risk of mortality is multiplied by 0.85.

###### 1.2.3.2 Protocolised $S/F_{94}$

In the ISARIC dataset, $S/F_{94}$ measurements are made opportunistically. We expect protocolised $S/F_{94}$ measurements to be more precise and to be a better predictor of mortality. This in turn implies that smaller sample sizes will be required when using protocolised $S/F_{94}$ measurements as an intermediate endpoint in clinical trials.

In order to calculate the magnitude of this effect, we assume a measurement error model of the following form

$$y_i = \alpha_0 + \alpha_1 x_i + \epsilon_i, \tag{1.8}$$

where $x_i$ is a protocolised measurement of $S/F_{94}$, $\alpha_0, \alpha_1$ are scalar coefficients, and $\epsilon_i$ is a residual.

We will use the notation

$$\mu_x := \mathbb{E}[x_i] \tag{1.9}$$
$$\mu_y := \mathbb{E}[y_i] \tag{1.10}$$
$$\sigma_x^2 := \mathrm{Var}[x_i] \tag{1.11}$$
$$\sigma_y^2 := \mathrm{Var}[y_i]. \tag{1.12}$$

We also assume the moment conditions

$$\mathbb{E}[\epsilon_i] = 0 \tag{1.13}$$
$$\mathbb{E}[\epsilon_i x_i] = 0. \tag{1.14}$$

Combining 1.13 and 1.14 with 1.8 gives

$$\alpha_0 = \mu_y - \frac{\rho_{xy}\sigma_y\mu_x}{\sigma_x} \tag{1.15}$$
$$\alpha_1 = \frac{\rho_{xy}\sigma_y}{\sigma_x}, \tag{1.16}$$

where $\rho_{xy}$ is the Pearson correlation between $x_i$ and $y_i$. Assuming $\{y_i\}_{i=1\ldots I}$ is mean and variance ergodic, we have the following consistent estimators for $\mu_y$ and $\sigma_y^2$

$$\hat{\mu}_y := \frac{1}{I}\sum_{i=1}^{I} y_i \tag{1.17}$$
$$\hat{\sigma}_y^2 := \frac{1}{I}\sum_{i=1}^{I}(y_i - \hat{\mu}_y)^2 \tag{1.18}$$

Combining 1.1 with 1.8 gives

$$\ln\left(\frac{p_i}{1-p_i}\right) = \beta_0' + \beta_1' x_i + \beta_1\epsilon_i + \beta \cdot z_i, \tag{1.19}$$

where $\beta_0' := \beta_0 + \alpha_0\beta_1$ and $\beta_1' = \alpha_1\beta_1$.

Given knowledge of $\mu_x$, $\sigma_x$ and $\rho_{xy}$, $\alpha_0$ and $\alpha_1$ can be consistently estimated by substituting 1.17, 1.18 into 1.15 and 1.16. Denoting these estimators $\hat{\alpha}_0$ and $\hat{\alpha}_1$ respectively, we define

$$\ln\left(\frac{\hat{p}_i}{1-\hat{p}_i}\right) := \hat{\beta}_0' + \hat{\beta}_1' x_i + \hat{\beta}\cdot z_i, \tag{1.20}$$

where $\hat{\beta}_0' := \hat{\beta}_0 + \hat{\alpha}_0\hat{\beta}_1$, $\hat{\beta}_1' = \hat{\alpha}_1\hat{\beta}_1$, and $\hat{p}_i$ is the corresponding estimate of the probability of day 28 mortality for individual $i$. We have

$$\hat{\beta}_0' \xrightarrow{p} \beta_0' \tag{1.21}$$
$$\hat{\beta}_1' \xrightarrow{p} \beta_1', \tag{1.22}$$

where $\xrightarrow{p}$ denotes convergence in probability. If in addition we assume $\{\epsilon_i\}_{i=1}^I$ is mean ergodic, then using 1.13 gives

$$\text{plim}_{I\to\infty}\frac{1}{I}\sum_{i=1}^{I}\left[\ln\left(\frac{p_i}{1-p_i}\right) - \ln\left(\frac{\hat{p}_i}{1-\hat{p}_i}\right)\right] = 0. \tag{1.23}$$

Now consider an individual-specific treatment effect given by

$$p_i \to f(p_i) \tag{1.24}$$

for some function f taking values in $[0,1]$, that may in principle depend on all covariates. Let $\Delta_i, \hat{\Delta}_i$ be the change in $x_i$ associated with this treatment effect in 1.19 and 1.20 respectively. That is,

$$\ln\left(\frac{f(p_i)}{1-f(p_i)}\right) = \beta_0' + \beta_1'(x_i+\Delta_i) + \beta_1\epsilon_i + \beta\cdot z_i. \tag{1.25}$$
$$\ln\left(\frac{f(\hat{p}_i)}{1-f(\hat{p}_i)}\right) = \hat{\beta}_0' + \hat{\beta}_1'(x_i+\hat{\Delta}_i) + \hat{\beta}\cdot z_i. \tag{1.26}$$

Our estimator for the average change in protocolised $S/F_{94}$ associated with the treatment effect $f$ is obtained by subtracting 1.20 from 1.26, dividing by $\hat{\beta}_1'$ and averaging over the sample,

$$\hat{\mu}_\Delta = \frac{1}{\hat{\beta}_1' I}\sum_{i=1}^{I}\ln\left(\frac{f(\hat{p}_i)(1-\hat{p}_i)}{\hat{p}_i(1-f(\hat{p}_i))}\right). \tag{1.27}$$

We now show this is a consistent estimator for $\mu_\Delta := \mathbb{E}[\Delta_i]$ at first order in $\epsilon_i$.

Consider

$$\hat{\mu}_\Delta - \frac{1}{I}\sum_{i=1}^{I}\Delta_i = \frac{1}{I}\sum_{i=1}^{I}\left(\frac{1}{\hat{\beta}_1'}\left[\ln\left(\frac{f(\hat{p}_i)}{1-f(\hat{p}_i)}\right) - \ln\left(\frac{\hat{p}_i}{1-\hat{p}_i}\right)\right] - \frac{1}{\beta_1'}\left[\ln\left(\frac{f(p_i)}{1-f(p_i)}\right) + \ln\left(\frac{p_i}{1-p_i}\right)\right]\right) \tag{1.28}$$

We assume $f$ is analytic, and Taylor expand the third term in 1.28 to first order around $\epsilon_i = 0$. Let $g(\epsilon_i) = \ln\left[\frac{f(p_i)}{1-f(p_i)}\right]$, where $p_i$ is understood to be a function of $\epsilon_i$ via 1.19. Then

$$\hat{\mu}_\Delta - \frac{1}{I}\sum_{i=1}^{I}\Delta_i \simeq \frac{1}{I}\sum_{i=1}^{I}\left(\frac{1}{\hat{\beta}_1'}\left[\ln\left(\frac{f(\hat{p}_i)}{1-f(\hat{p}_i)}\right) - \ln\left(\frac{\hat{p}_i}{1-\hat{p}_i}\right)\right]\right.$$
$$\left. - \frac{1}{\beta_1'}\left[g(\epsilon_i = 0) + g'(\epsilon_i = 0)\epsilon_i + \ln\left(\frac{p_i}{1-p_i}\right)\right]\right) \tag{1.29}$$

Now taking probability limits and using 1.22, 1.23, as well as

$$\text{plim}_{I\to\infty}\left(g(\epsilon_i = 0) - \ln\left(\frac{f(\hat{p}_i)}{1-f(\hat{p}_i)}\right)\right) = 0 \tag{1.30}$$
$$\text{plim}_{I\to\infty}\sum_{i=1}^{I}\epsilon_i = 0, \tag{1.31}$$

we obtain

$$\text{plim}_{I\to\infty}\left(\hat{\mu}_\Delta - \mu_\Delta\right) = 0, \tag{1.32}$$

where we have assumed $\{\Delta_i\}_{i=1}^{I}$ is mean ergodic. This is the desired consistency result.

Note that the form of the measurement error model 1.8 places some restriction on feasible values for $\rho_{xy}$. If we denote opportunistic and protocolised measurements on day $d$ by $y_d, x_d$ respectively, then using 1.8 and assuming $\sigma_x, \sigma_y$ are constant over time we obtain

$$\text{Cov}(y_0, y_5) = \alpha_1^2\text{Cov}(x_0, x_5). \tag{1.33}$$

Substituting in (5.16) gives

$$\text{Corr}(y_0, y_5) = \rho_{xy}^2\text{Corr}(x_0, x_5), \tag{1.34}$$

where Corr(.,.) denotes the pearson correlation. Since the pearson correlation takes values in $[0,1]$, we have

$$\text{Corr}(y_0, y_5) \in [0, \rho_{xy}^2]. \tag{1.35}$$

We calculated effect sizes with protocolised $S/F_{94}$ as outcome measure, with the means of the opportunistic and protocolised measurements set equal, and the standard deviation of protocolised $S/F_{94}$ measurements set to 80% of the standard deviation of opportunistic SF/94 measurements. In the ISARIC4C data, the sample correlation between opportunistic $S/F_{94}$ measurements on day 0 and day 5 was $\simeq 0.40$. Using 1.36 gives the constraint

$$\rho_{xy} \geq 0.63. \tag{1.36}$$

###### 1.2.3.3 WHO ordinal scale

We used a proportional odds model to estimate the relationship between day 28 WHO ordinal scale category and individual characteristics,

$$\ln\left(\frac{p_{ij}}{1-p_{ij}}\right) = \beta_{0j} + \beta \cdot z_i. \tag{1.37}$$

Here, $p_{ij}$ is the probability that the WHO ordinal scale category of individual $i$ at day 28 is $\leq j$, $\beta_{0j}$ are a set of constant terms for each WHO ordinal scale category, $\beta$ is an $n$-dimensional vector of coefficients, and $z_i$ is an $n$-dimensional vector of covariates.

$j = 10$ is the highest WHO ordinal scale category and corresponds to death. Then $q_{i10} := 1 - p_{i10}$ is the probability of day 28 mortality for patient $i$. We consider an individual-specific treatment effect given by

$$q_{i10} \to f(q_{i10}) \tag{1.38}$$

for some function f taking values in $[0, 1]$, that may in principle depend on all covariates.

Let $\hat{q}_{i10}$ denote the predicted probability of day 28 mortality for individual $i$ derived from the model 1.37. Then the rate of predicted day 28 mortality in the sample can be estimated by $\bar{q}_{10} := \frac{1}{I}\sum_{i=1}^{I} \hat{q}_{i10}$. The odds ratio associated with the treatment effect $f$ is estimated by

$$\frac{\left(\frac{f(\bar{q}_{10})}{1-f(\bar{q}_{10})}\right)}{\left(\frac{\bar{q}_{10}}{1-\bar{q}_{10}}\right)} = \frac{f(\bar{q}_{10})\,(1-\bar{q}_{10})}{\bar{q}_{10}[1-f(\bar{q}_{10})]}. \tag{1.39}$$

We take this as our estimate of effect size.

##### 1.2.4 Generation of synthetic data

A synthetic dataset was generated for use in the online sample size calculation tool. This dataset was achieved using imputation by chained equations in multiple stages. First, empty records for one thousand new patients at day zero were added to the real data. Next, variables that can be reasonably expected to be constant over a 28 day time period (e.g comorbidities such as obesity, dementia, liver disease etc) were imputed by chained equations. New rows were added for day 5 and day 8 for each simulated record, with the constant variables imputed in the previous step repeated. After this, there were multiple rounds of partitioning the full dataset, containing both real and simulated records, and carrying out imputation by chained equations separately within each piece. For example, a binary variable indicating whether an individual died was imputed, then the full dataset was split into those who died and those who didn't, and a day of death was imputed for the former. Similarly, binary variables indicating whether or not the individual had a one/two level sustained improvement in WHO ordinal severity score were imputed, and used to partition the full dataset. Then the minimum time taken for patients to have a sustained one/two level improvement was imputed amongst those that had this event occur. Appropriate care was taken in order to respect variable bounds, interdependence and the time series structure of the data. For example, if an individual had a sustained two level improvement, they necessarily must also have had a sustained one level improvement, and the time taken for the former to occur must necessarily be greater than or equal to the time taken for the latter to occur. In order to handle this appropriately, we first imputed the time taken for a sustained one level improvement, and after appropriate partitioning, we then imputed the difference in time taken for a sustained one and two level improvement using predictive mean matching. This guaranteed a positive difference, as required.

In each imputation by chained equations, predictive mean matching was used for continuous variables and logistic or multinomial logistic regression was used for categorical variables.

#### 1.3 Inclusion criteria

We compared the effect on sample size using different inclusion criteria (Supplementary Tables 2, 3 and 4). In general, as expected, the more broad the inclusion criteria (e.g. only age limits), the larger the sample size that is needed; and the more specific the inclusion criteria, the smaller the sample size. However it is important to note that using very specific inclusion criteria increases the burden associated with screening patients for inclusion in a study, which may substantially decrease real-world recruitment.

| Selection criteria | $S_aO_2 < 0.94$ or $F_IO_2 = 0.21$ | $S_aO_2 > 0.94$ | Day 0 and ($S_aO_2 < 0.94$ or $F_IO_2 = 0.21$) | Day 0 and $S_aO_2 > 0.94$ |
|---|---|---|---|---|
| Number of entries | 192,583 | 55,765 | 114,668 | 27,367 |
| Unique subjects | 129,049 | 39,457 | 114,688 | 27,367 |
| Age, years | 71 (57-82) | 69 (55-82) | 72 (58-83) | 70 (55-82) |
| Male | 111,244 (58%) | 32,022 (58%) | 63,127 (55%) | 15,351 (56%) |
| Respiratory rate, per minute | 20 (18-25) | 22 (18-26) | 20 (18-25) | 22 (18-25) |
| Temperature, Celsius | 37.2 (36.8-38.0) | 37.1 (36.7-37.8) | 37.3 (36.8-38.1) | 37.3 (36.8-38.1) |
| Systolic blood pressure, mmHg | 116 (103-138) | 120 (105-140) | 117 (104-138) | 121 (106-142) |
| Diastolic blood pressure, mmHg | 66 (57-77) | 69 (59-77) | 68 (58-79) | 70 (60-81) |
| CRP, mg/L | 85 (39-159) | 84 (39-152) | 78 (34-145) | 84 (40-149) |
| BUN, mmol/L | 7.0 (4.8-10.9) | 6.7 (4.6-10.5) | 6.7 (4.6-10.3) | 6.5 (4.5-10.1) |
| Creatinine, umol/L | 86 (68-120) | 84 (67-119) | 88 (70-119) | 86 (70-116) |
| Urine production, ml/24h | 1,001 (400-1839) | 1,020 (450-1890) | 500 (300-950) | 550 (300-1,000) |
| Glasgow Coma Scale | 15 (15-15) | 15 (15-15) | 15 (15-15) | 15 (15-15) |
| Clinical frailty | 4 (2-6) | 4 (2-6) | 4 (3-6) | 4 (2-6) |
| Onset to admission, days | 4 (0-8) | 4 (0-8) | 3 (0-7) | 4 (0-8) |
| Infiltrates on X-ray present | 52,001 (67%) | 14,525 (69%) | 41,944 (63%) | 12,147 (67%) |
| No respiratory support (WHO 4) | 71,633 (37%) | 0 (0%) | 11,187 (23%) | 0 (0%) |
| Nasal oxygen (WHO 5) | 53,610 (28%) | 23,220 (42%) | 19,904 (41%) | 10,102 (64%) |
| Non invasive ventilation (WHO 6) | 46,473 (24%) | 11,764 (21%) | 15,370 (32%) | 5,148 (32%) |
| IMV: SF >2.0 (WHO 7) | 3,063 (2%) | 1,789 (3%) | 249 (<1%) | 172 (1%) |
| IMV: SF<2.0 or vasopressor (WHO 8) | 8,782 (5%) | 1,900 (3%) | 1,174 (2%) | 345 (2%) |
| IMV: SF<2.0 and vasopressor, ECMO or dialysis (WHO 9) | 3,004 (2%) | 784 (1%) | 401 (<1%) | 151 (1%) |
| Death (WHO 10) | 30,225 (23%) | 8,289 (21%) | 26,017 (23%) | 5,924 (22%) |
| Discharged alive (including transfer to other facility) | 98,824 (77%) | 31,168 (79%) | 88,651 (77%) | 21,443 (78%) |

Supplementary Table 1: Characteristics of patient populations meeting criteria for $S/F$ 94 measurement in ISARIC4C data. Continuous values are summarised as median (inter quartile range). Categorical values are summarised as frequency (percentage). Subjects meeting the criteria for $S/F$ 94 ($S_aO_2 < 0.94$ or $F_IO_2 = 0.21$) were compared to patients not meeting the $S/F$ 94 criteria. Comparison was done both for all entries during admission and day 0. CRP = C-reactive protein, BUN= blood urea nitrogen

| Measure | Distribution/ Event rate | Estimated treatment effect | Total $n$ required 80% power $2p = 0.05$ |
|---|---|---|---|
| $S/F_{94}$ day 5 | Mean = 2.40<br>SD = 1.32<br>$\rho$ vs Day 0: 0.43 | Difference in $S/F_{94}$: 0.175 | 1,454 |
| $S/F_{94}$ day 8 | Mean = 2.74<br>SD = 1.54<br>$\rho$ vs Day 0: 0.40 | Difference in $S/F_{94}$: 0.154 | 2,636 |
| WHO day 5 | 4: 1,502/8,315<br>5: 1,861/8,315<br>6: 2,270/8,315<br>7: 301/8,315<br>8: 980/8,315<br>9: 313/8,315<br>10: 1,088/8,315 | Proportional odds ratio: 0.83 | 2,881 |
| WHO day 8 | 4: 1,436/6,942<br>5: 1,206/6,942<br>6: 1,196/6,942<br>7: 361/6,942<br>8: 812/6,942<br>9: 261/6,942<br>10: 1,670/6,942 | Proportional odds ratio: 0.81 | 2,234 |
| 1-level sustained improvement | 12,057/26,854 (44.9%) | Risk ratio: 1.03 | 6,544 |
| 2-level sustained improvement | 4,914/26,854 (18.3%) | Risk ratio: 1.04 | 3,722 |
| 28-day mortality | 7,583/36,559 (20.7%) | Risk ratio: 0.85 | 5,154 |

Supplementary Table 2: Comparison of outcome measures among 36,559 hospitalised patients aged 20-75, who required supplemental oxygen on the day of admission. The estimated treatment effect is for a 15% relative reduction in mortality. For $S/F_{94}$, the mean value and standard deviation (SD) and the correlation with $S/F_{94}$ on day 0 are reported for both day 5 and day 8. For the WHO ordinal severity scale, the number of people in each group between 4 (hospitalised, no oxygen support) and 10 (dead) are reported. For 1- and 2-level sustained improvement, the number of people reaching sustained improvement is reported. For 28-day mortality, the proportion of patients who died at day 28 is reported. Sample sizes shows the total number of subjects needed in both arms using a 1:1 allocation.

| Measure | Distribution/ Event rate | Estimated treatment effect | Total $n$ required 80% power $2p = 0.05$ |
|---|---|---|---|
| $S/F_{94}$ day 5 | Mean = 2.84<br>SD = 1.47<br>$\rho$ vs Day 0: 0.56 | Difference in $S/F_{94}$: 0.172 | 1,566 |
| $S/F_{94}$ day 8 | Mean = 3.15<br>SD = 1.57<br>$\rho$ vs Day 0: 0.47 | Difference in $S/F_{94}$: 0.151 | 2,650 |
| WHO day 5 | 4: 3,162/14,082<br>5: 2,903/14,082<br>6: 3,091/14,082<br>7: 385/14,082<br>8: 1,198/14,082<br>9: 438/14,082<br>10: 2,905/14,082 | Proportional odds ratio: 0.82 | 2,431 |
| WHO day 8 | 4: 2,708/11,953<br>5: 1,880/11,953<br>6: 1,688/11,953<br>7: 455/11,953<br>8: 1,037/11,953<br>9: 370/11,953<br>10: 3,815/11,953 | Proportional odds ratio: 0.79 | 1,855 |
| 1-level sustained improvement | 14,556/47,260 (30.8%) | Risk ratio: 1.03 | 6,061 |
| 2-level sustained improvement | 5,860/47,260 (12.4%) | Risk ratio: 1.04 | 2,959 |
| 28-day mortality | 10,811/69,464 (15.6%) | Risk ratio: 0.85 | 7,267 |

Supplementary Table 3: Comparison of outcome measures among 69,464 hospitalised patients aged 20-75. The estimated treatment effect is for a 15% relative reduction in mortality. For $S/F_{94}$, the mean value and standard deviation (SD) and the correlation with $S/F_{94}$ on day 0 are reported for both day 5 and day 8. For the WHO ordinal severity scale, the number of people in each group between 4 (hospitalised, no oxygen support) and 10 (dead) are reported. For 1- and 2-level sustained improvement, the number of people reaching sustained improvement is reported. For 28-day mortality, the proportion of patients who died at day 28 is reported. Sample sizes shows the total number of subjects needed in both arms using a 1:1 allocation.

| Measure | Distribution/ Event rate | Estimated treatment effect | Total $n$ required 80% power $2p = 0.05$ |
|---|---|---|---|
| $S/F_{94}$ day 5 | Mean = 2.40<br>SD = 1.29<br>$\rho$ vs Day 0: 0.32 | Difference in $S/F_{94}$: 0.180 | 1,462 |
| $S/F_{94}$ day 8 | Mean = 2.74<br>SD = 1.51<br>$\rho$ vs Day 0: 0.28 | Difference in $S/F_{94}$: 0.158 | 2,644 |
| WHO day 5 | 4: 1,770/10,039<br>5: 2,433/10,039<br>6: 2,736/10,039<br>7: 363/10,039<br>8: 1,145/10,039<br>9: 418/10,039<br>10: 1,174/10,039 | Proportional odds ratio: 0.83 | 2,971 |
| WHO day 8 | 4: 1,685/8,240<br>5: 1,525/8,240<br>6: 1,445/8,240<br>7: 429/8,240<br>8: 968/8,240<br>9: 343/8,240<br>10: 1,844/8,240 | Proportional odds ratio: 0.81 | 2,321 |
| 1-level sustained improvement | 13,437/30,060 (44.7%) | Risk ratio: 1.03 | 6,756 |
| 2-level sustained improvement | 5,411/30,060 (18.0%) | Risk ratio: 1.04 | 3,808 |
| 28-day mortality | 8,262/39,765 (20.8%) | Risk ratio: 0.85 | 5,143 |

Supplementary Table 4: Comparison of outcome measures among 39,765 hospitalised patients aged 20-75, who required supplemental oxygen in the first 3 days in hospital. The estimated treatment effect is for a 15% relative reduction in mortality
